# Proteomic blood profiling in mild, severe and critical COVID-19 patients

**DOI:** 10.1101/2020.06.22.20137216

**Authors:** Hamel Patel, Nicholas J Ashton, Richard J.B Dobson, Lars-Magnus Andersson, Aylin Yilmaz, Kaj Blennow, Magnus Gisslen, Henrik Zetterberg

## Abstract

The recent SARS-CoV-2 pandemic manifests itself as a mild respiratory tract infection in the majority of individuals leading to COVID-19 disease. However, in some infected individuals, this can progress to severe pneumonia and acute respiratory distress syndrome (ARDS), leading to multi-organ failure and death. The purpose of this study is to explore the proteomic differences between mild, severe and critical COVID-19 positive patients. Blood protein profiling was performed on 59 COVID-19 mild (n=26), severe (n=9) or critical (n=24) cases and 28 controls using the OLINK inflammation, autoimmune, cardiovascular and neurology panels. Differential expression analysis was performed within and between disease groups to generate nine different analyses. From the 368 proteins measured per individual, more than 75% were observed to be significantly perturbed in COVID-19 cases. Six proteins (IL6, CKAP4, Gal-9, IL-1ra, LILRB4 and PD-L1) were identified to be associated with disease severity. The results have been made readily available through an interactive web-based application for instant data exploration and visualization, and can be accessed at https://phidatalab-shiny.rosalind.kcl.ac.uk/COVID19/. Our results demonstrate that dynamic changes in blood proteins that associate with disease severity can potentially be used as early biomarkers to monitor disease severity in COVID-19 and serve as potential therapeutic targets.

## Introduction

The recent severe acute respiratory syndrome coronavirus 2 (SARS-CoV-2) pandemic manifests itself as a mild respiratory tract infection in the majority of individuals leading to coronavirus disease 2019 (COVID-19) disease. However, in some infected individuals, this can progress to severe pneumonia and acute respiratory distress syndrome (ARDS), leading to multi-organ failure and death. The exact percentage of patients presenting with severe symptoms is currently impossible to calculate as the exact number of individuals who have contracted the virus is unknown and many who have, are unaware due to being asymptomatic. Nevertheless, according to the World Health Organization (WHO), it is estimated 80% of infections are asymptomatic or mild, 15% are severe infections requiring oxygen support, and 5% are critical requiring ventilation (1).

SARS-CoV-2 shares an evolutionary relationship with SARS-CoV, the causative pathogen of the SARS outbreak in 2013 (2). Currently, owing to the lack of reliable markers, it is challenging to monitor individuals that are progressing to severe COVID-19, which relies mainly on clinical manifestations (3). Previous studies on highly pathogenic coronaviruses, e.g., SARS or Middle East Respiratory Syndrome (MERS), have highlighted an inflammatory cytokine storm and lymphocytopenia as common features relating to disease severity (4–6). It has also been suggested that the presenting cytokine storm is related to rapid disease progression or inadequate response to treatment (6). Thus, it is vitally essential to tease out which peripheral markers are reliably related to disease severity to administer treatment at the earliest stage.

Furthermore, central nervous system (CNS) involvement has also been reported in hospitalized patients infected with SARS (7). It is plausible that patients suffering from COVID-19 might also exhibit CNS damage. Our previous results show neurochemical evidence of neuronal injury and glial response in patients with severe and critical COVID-19 and are associated with disease severity (8). Therefore, it is plausible that other CNS injury markers can be detected in the blood to support the possible impact of COVID-19 on the CNS. In this study, by examining an extensive collection of inflammatory, immune response, cardiovascular and neurological markers in the blood, we hope to highlight biomarkers, which demonstrate the progression from mild to critical disease.

## Results

### Cohort demographics

An overview of the patient demographics is provided in Table 1 and significance testing of age, gender and “*days since symptom onset*” between groups is provided in Supplementary Table 1. The mild group was identified to contain patients significantly younger when compared to the remaining groups (control p-value = 0.01, severe p-value = 0.02 and critical p-value = 0.01). No significant difference in age was identified between the remainder of the groups, including between the control group and the case group (p-value = 0.15).

**Table 1:** Cohort demographics

|  |  | Control | Mild | Severe | Critical | Total cases |
| --- | --- | --- | --- | --- | --- | --- |
| Participants (n) |  | 28 | 26 | 9 | 24 | 59 |
| Follow-up (n) |  | 0 | 6 | 0 | 5 | 11 |
| Gender (M/F) |  | 15/13 | 13/13 | 6/3 | 22/2 | 41/18 |
| Age (95% CI) |  | 63.1 (55.9-70.3) | 51.3 (45.9 - 56.8) | 64.8 (55.0 - 74.6) | 61.0 (56.2-65.8) | 57.8 (53.8 – 60.8) |
| Ethnicity (n) | African black | NA | 0 | 0 | 1 | 1 |
|  | Caucasian | NA | 20 | 8 | 23 | 51 |
|  | Persian | NA | 6 | 1 | 0 | 7 |
| “Days since symptom onset” (95% CI) |  | NA | 20.9 (17.3 - 24.5) | 12 (10.3 - 13.7) | 11 (9.1-13) | 15.5 (13.4-17.7) |

The table provides a summary of the available demographics in this study. Cases represent the mild, the severe, and the critical group merged into one group. The “Follow-up” represents the number of patients that had samples taken at two different time points. The mild group was identified to contain patients significantly younger when compared to the remaining groups (control $p$ -value = 0.01, severe $p$ -value = 0.02 and the critical $p$ -value = 0.01). Gender was identified to be significantly different between the control and the critical group ( $p$ -value = 0.005) and between the mild and the critical group ( $p$ -value = 0.002) only. The number of “days since symptom onset” was found to be significantly higher in the mild group when compared to the severe ( $p$ -value = $5.41e-05$ ) and the critical group ( $p$ -value = $1.60e-05$ ). Ethnicity information was unavailable for the control group. Abbreviations: M = Males, F = Females, CI = Confidence Interval.

Gender was identified to be significantly different between the control and the critical group (p-value = 0.005) and between the mild and the critical group (p-value = 0.002) only. There was no significant difference in gender distribution between the control group and the case group (p-value = 0.16). The number “*days since symptom onset*” was found to be significantly higher in the mild group when compared to the severe (p-value 5.41e-05) and critical group (p-value = 1.60e-05), with no difference between the severe and the critical group (p-value = 0.434).

### Summary of OLINK data processing

The OLINK cardiovascular, immune, inflammation and neurology panels consisted of 92 proteins each. Protein profiling the 87 samples resulted in the measurement of 368 proteins per sample. One sample, belonging to the mild symptom group, failed in two assays (immune and neurology) and was excluded from the failed panels, but were retained for analysis involving the cardiovascular and inflammation panels.

Thirteen proteins had missing Normalized Protein eXpression (NPX) values or had NPX values below the protein-specific limit of detection (LOD) in more than 50% of samples in all four disease groups. These 13 proteins were removed, leaving 355 proteins, of which 344 proteins were unique due to protein duplication across the four panels.

### Summary of differential expression analysis

A summary of the number of proteins being significantly perturbed in each analysis is provided in Table 2. The complete differential expression (DE) results for all analyses are available in Supplementary Table 2 and can also be explored using the data explorer application developed in this study (https://phidatalab-shiny.rosalind.kcl.ac.uk/COVID19/).

**Table 2:** Summary of differential expression analysis

| Analysis | Total DE | Up | Down | Most sig | logFC | FDR p-value |
| --- | --- | --- | --- | --- | --- | --- |
| control vs mild | 147 | 42 | 105 | NF2 | -<br>4.62E+00 | 2.57E-42 |
| control vs severe | 65 | 8 | 57 | MANF | -<br>4.71E+00 | 5.12E-16 |
| control vs critical | 187 | 82 | 105 | NF2 | -<br>4.44E+00 | 2.40E-36 |
| mild vs severe | 146 | 97 | 49 | EN-RAGE | 2.16E+00 | 5.33E-09 |
| mild vs critical | 197 | 158 | 39 | CTSL1 | 2.16E+00 | 3.76E-13 |
| severe vs critical | 64 | 63 | 1 | IL-18R1 | 5.79E-01 | 0.00139 |
| control vs case | 269 | 120 | 149 | NF2 | -<br>4.62E+00 | 1.31E-86 |
| mild group longitudinal | 13 | 10 | 3 | BOC | 1.57E+00 | 0.001243 |
| critical group longitudinal | 6 | 3 | 3 | DECR1 | 0.6896971 | 0.023089 |

The table provides a summary of the number of significantly ( $FDR\ p\text{-value} \leq 0.05$ ) differentially expressed proteins identified in all nine analyses. “Total DE” is the total number of differentially expressed proteins identified in the analysis, “Up” represents the number of proteins that are up-regulated and “Down” represents the number of down-regulated proteins. “Most sig” represents the most significant perturbed protein in the analysis and is provided as the protein symbol. Abbreviations: logFC = log fold change, FDR = false discovery rate.

### Proteins differentially expressed in COVID-19

The “control vs case” analysis identified 269 proteins as significantly differentially expressed in COVID-19 patients, with the NF2 protein identified as the most perturbed (FDR p-value = 1.31E-86, logFC = −4.62E+00). The NF2 protein was also identified as the most perturbed protein in multiple analyses (“control vs mild”, “control vs critical” and “control vs case”) and was the single most perturbed protein from all nine analyses. The expression pattern of the NF2 protein per sample is illustrated in Figure 1.

**Figure 1:**
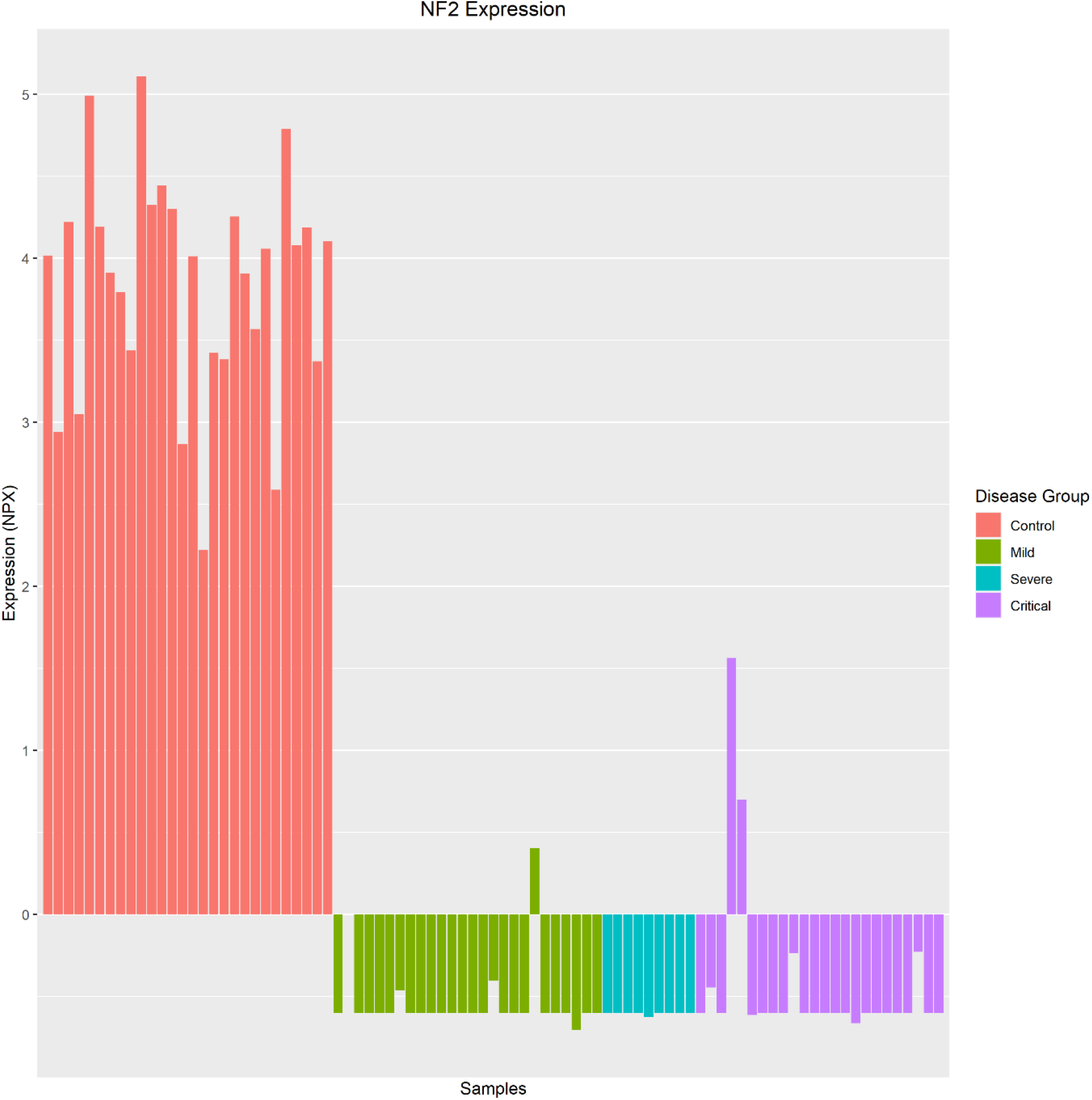
NF2 Expression across all disease groups. This protein was identified as the most perturbed protein in the “control vs case” analysis and is significantly down-regulated in all COVID-19 patients, regardless of infection severity.

The 269 proteins perturbed in COVID-19 mapped to 265 unique Entrez gene IDs in the ConsensusPathDB database, which identified 285 significantly enriched biological pathways. The most significantly enriched pathway was the “Cytokine-cytokine receptor interaction” (FDR adjusted p-value = 1.72 e-41). The pathway, along with the enriched proteins, is illustrated in Figure 1Figure 2. The complete pathway enrichment analysis results are provided in Supplementary Table 3

**Figure 2:**
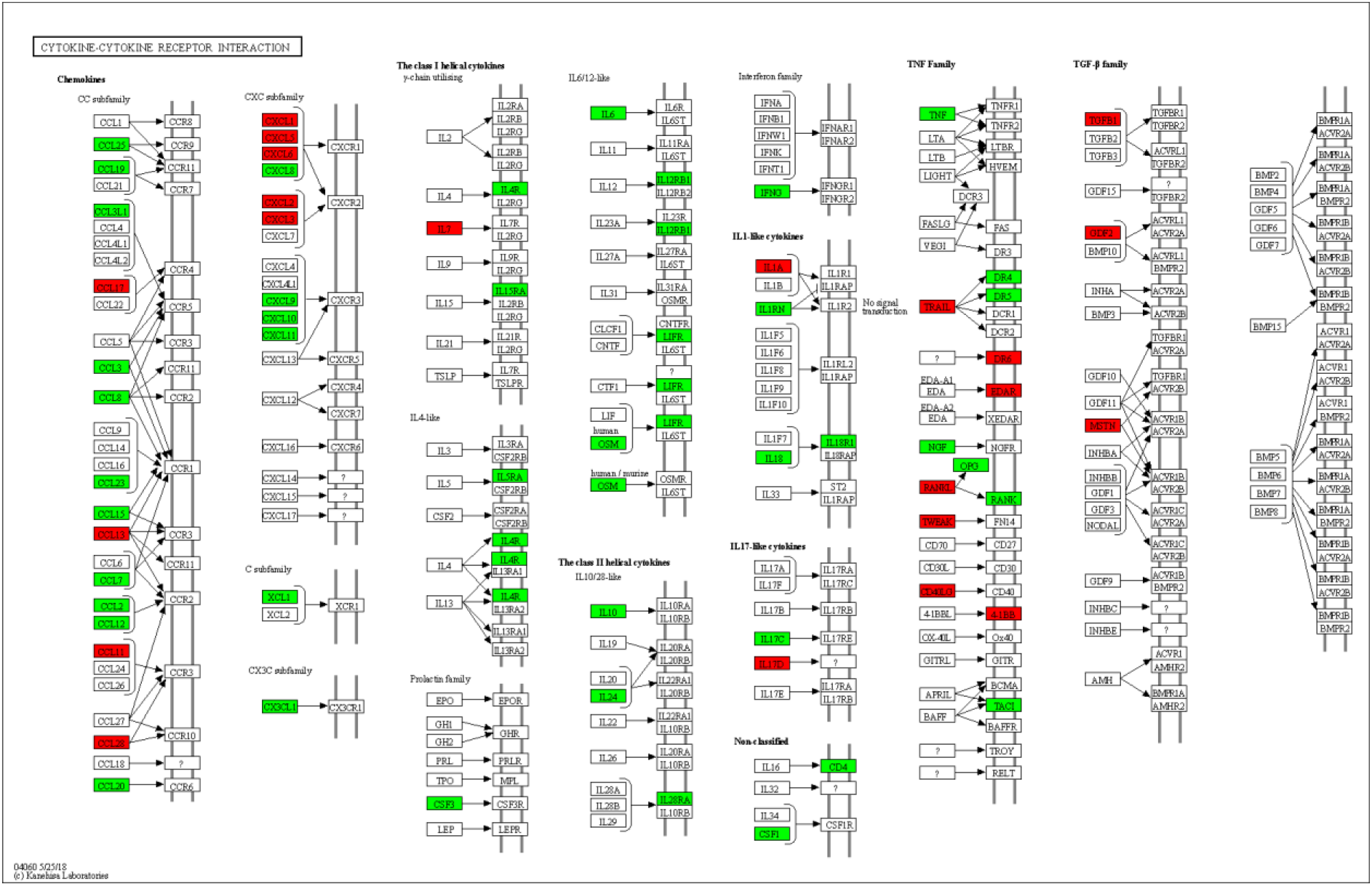
Cytokine-cytokine receptor interaction. The 269 proteins significantly differentially expressed in COVID-19 patients were significantly enriched in the “cytokine-cytokine receptor interaction” pathway, which is illustrated above. The proteins highlighted in green are up-regulated, and the proteins highlighted in red were down-regulated in COVID-19. The figure was generated using the “KEGG mapper – Search&Color Pathway” (Kanehisa & Sato, 2020).

### Proteins associated with COVID-19 severity

Eleven proteins were identified to be significantly differentially expressed between the control, the mild, the severe and the critical symptom groups. Eight of these proteins were consistently perturbed in the same direction as the infection symptoms increased (control −> mild −> severe −> critical). Due to the presence of the duplicate proteins across the different panels, these eight proteins correspond to six unique proteins. The six proteins are IL6, CKAP4, Gal-9, IL-1ra, LILRB4 and PD-L1, and are deemed to be associated with COVID-19 severity. None of these proteins originates from the neurology panel and their statistical significance across the nine analyses is included in Supplementary Table 2.

The IL6 protein was discovered to have been repeated on three different panels, and its expression pattern and significance level were found to be very similar between the disease groups. When comparing the overall magnitude of expression change from the control group to the critical group (“control vs critical” analysis), IL6 protein on the immune, inflammation and the cardiovascular panel has a logFC of 4.23, 4.49, 4.79, and an FDR p-value of 4.01e-10, 2.74e-10 and 2.61e-10, respectively. The consistency in results provides validity across the different panels and demonstrates that IL6 protein is reliably detected as significantly increased in COVID-19.

The expression of all six proteins was observed to significantly increase from the control group to the mild group, which then further increases in the severe group, and increases even further in the critical group. Their expression patterns for these proteins is shown in Figure 3: Six proteins a) IL6, b) PD-L1, c) CKAP4, d) IL-1ra e) Gal-9 and f) LILRB4 are consistently differentially expressed between the control, the mild, the severe and the critical symptom groups after controlling for age, gender, and “days since symptom onset”, suggesting these proteins may be associated with disease severity.. The proteins can be ranked by their magnitude of fold change from the control group to the critical group (as determined from the”control vs critical” analysis) as follows; IL6 (logFC=4.79), IL-1ra (logFC=2.43), CKAP4 (logFC=1.98), LILRB4 (logFC=1.79), Gal-9 (logFC=1.60), PD-L1 (logFC=1.25). Pathway analysis identified all six proteins as significantly enriched in the “Immune system” (FDR adjusted p-value = 1e-4).

**Figure 3:**
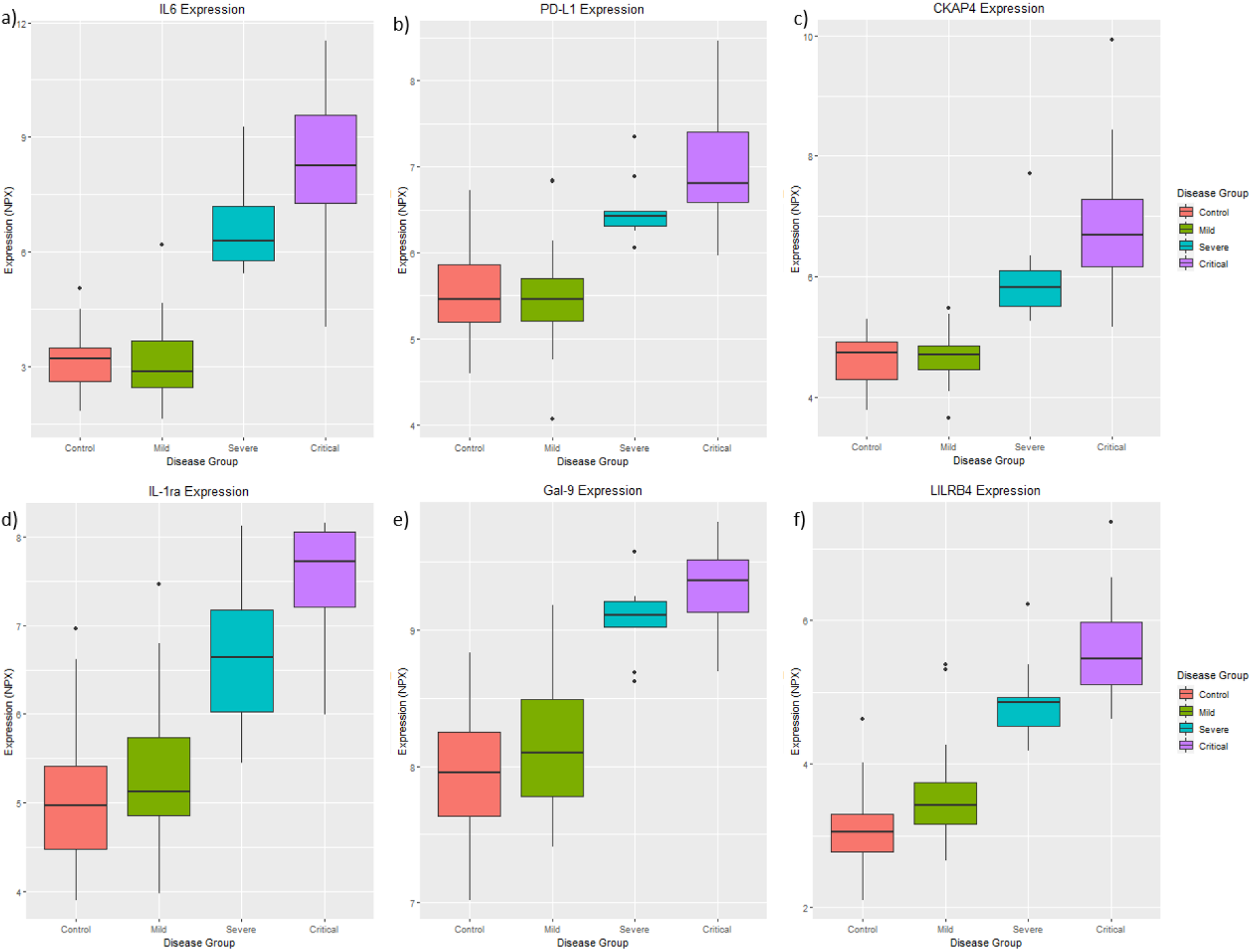
Six proteins a) IL6, b) PD-L1, c) CKAP4, d) IL-1ra e) Gal-9 and f) LILRB4 are consistently differentially expressed between the control, the mild, the severe and the critical symptom groups after controlling for age, gender, and “days since symptom onset”, suggesting these proteins may be associated with disease severity.

### Longitudinal data analysis

The mild symptom group (six patients) and the critical symptom group (five patients) consisted of patients where blood samples were taken at two different time points after the onset of disease symptoms. A summary of the demographics of these longitudinal samples is provided in Table 3: Demographics of the longitudinal samples. A summary of the number of proteins perturbed in each longitudinal analysis is included in Table 2, and the complete DE analysis results from these analyses are included in Supplementary Table 2. No protein was identified to be significantly perturbed in both the “mild group longitudinal” and the “critical group longitudinal” analyses.

**Table 3:** Demographics of the longitudinal samples

|  |  | Mild | Critical |
| --- | --- | --- | --- |
| No. patients |  | 6 | 5 |
| Gender (M/F) |  | 3/3 | 5/0 |
| Age (95% CI) |  | 51.5 (36.8 - 66.2) | 55.6 (45.8 - 73.4) |
| Ethnicity | Caucasian | 4 | 5 |
|  | Persian | 2 | 0 |
| Baseline sample symptom duration (95% CI) |  | 8.5 (3.4-13.6) | 8.4 (5.5 - 11.3) |
| Repeat sample symptom duration (95% CI) |  | 24.7 (22.2 - 27.1) | 11.2 (7.33 - 15.1) |
| Average time between samples (95% CI) |  | 16.2 (12.1 - 20.2) | 2.8 (1.4 - 4.2) |

The table provides the demographics of the longitudinal samples in this study. Abbreviations: M = Males, F = Females, CI = Confidence Interval.

### OLINK neuronal proteins correlated with markers of neural injury and astrogliosis

The three proteins measured on the Simoa platform were absent from the OLINK platform. The three proteins tau, NfL and GFAP were significantly (p-value ≤ 0.05) correlated with 97, 233, 165 proteins from the OLINK platform, respectively (Supplementary Table 4), of which 20, 61 and 41 proteins belong to the Neurology panel, respectively.

NfL was identified to be most correlated with EDA2R (r=0.66, p-value=4.01e-12), which is not significantly perturbed in any of the nine DE analysis performed in this study (see Figure 4a). Tau and GFAP were identified to be both most correlated with SCARB2 (tau: r=0.39 and p-value=1.74e-4, GFAP: r=0.46, p-value=6.47e-6). When compared to controls, SCARB2 is significantly up-regulated in the mild (logFC=0.59, p-value=4.7e-3) and critical groups (logFC=0.72, p-value=0.01) but not in the severe group (logFC=1.02, p-value=0.12). Although SCRAB2 is not statistically associated with disease severity, the expression of this protein is noted to increase inline with disease severity (Figure 4b).

**Figure 4:**
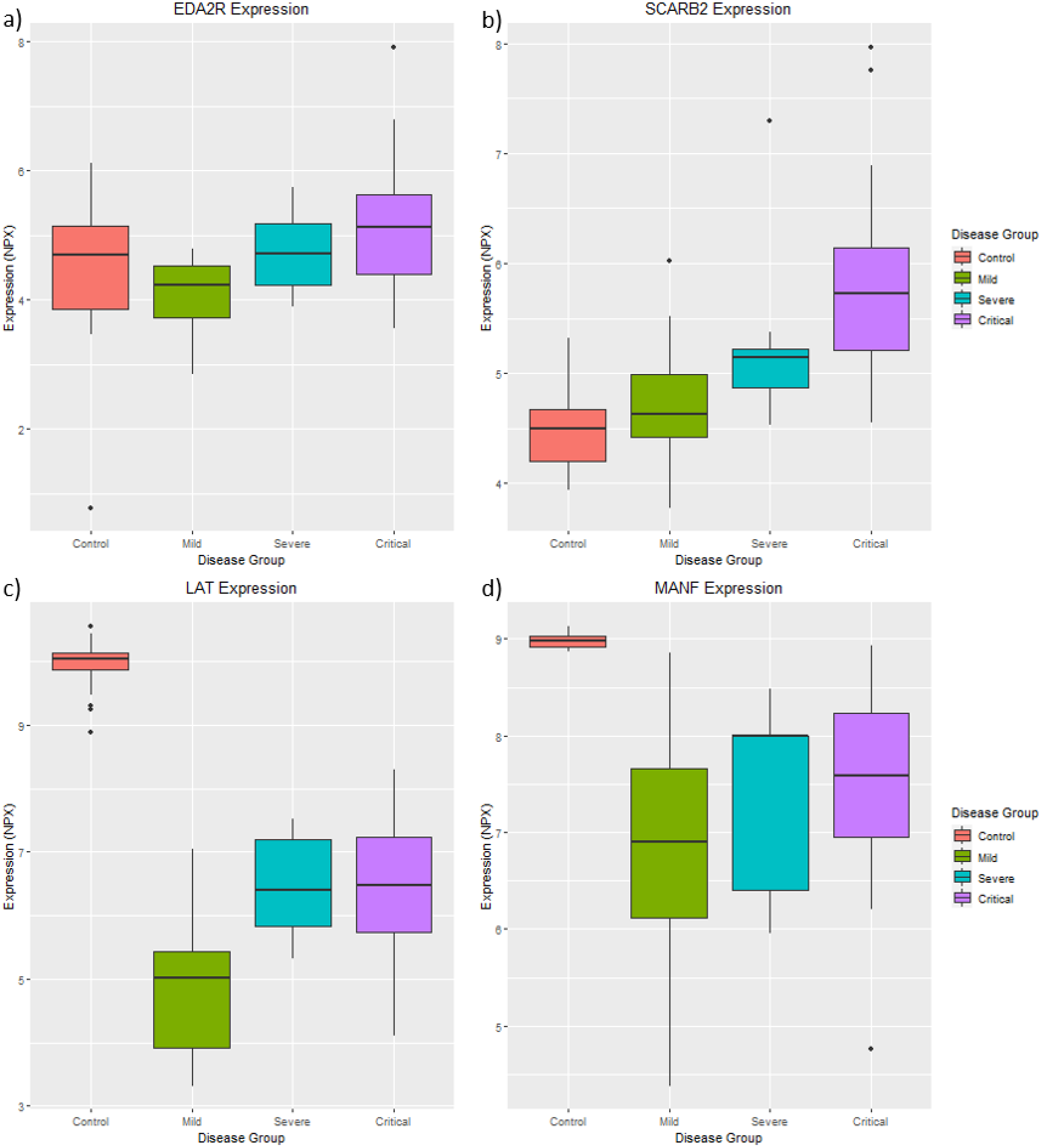
Expression patterns of the neuronal proteins perturbed in COVID-19. A) EDA2R is correlated with the Simoa measured NfL. B) SCARB2 is correlated with the Simoa measured tau and GFAP. C) LAT is most differentially expressed protein in the “control vs case” analysis. D) MANF is the most differentially expressed neuronal protein in this study from all analyses performed in this study. Statistical analysis was performed while controlling for age, gender, and “days since symptom onset.”

In addition, biomarkers of disease severity (Figure 3) were vastly more associated with NfL (CKAP4, r=0.642; PD-L1, r=0-623; IL6, r=0.528; Gal-9, LILRB4, r=0.508; r=0.486; IL-1ra, r=0.435) than GFAP or tau indicating that disease severity includes increased CNS damage as marked by NfL and not GFAP and tau.

### Differentially expressed OLINK neuronal proteins

From the “control vs case” analysis, 71 proteins on the neurology panel were identified to be perturbed in COVID-19, with LAT (see Figure 4c) identified to be the most perturbed in this analysis (logFC=-3.9, p-value=4.46e-22). LAT is also significantly differentially expressed between the mild and severe group (logFC=1.94, p-value=7.7e-3), but not between the severe and critical group (logFC=-0.08, p-value=0.89). This suggests LAT is down-regulated in COVID-19 but is not associated with disease severity. From all analyses involving the neuronal panel, MANF (see Figure 4d) was the most perturbed protein, and is significantly down-regulated in the mild (logFC= −2.85, p-value= 2.53e-28), severe (logFC=-4.71, p-value=8.58) and critical symptom groups (logFC=-1.81, p-value=5.98e-17). However, MANF is not significantly perturbed between the mild and severe group (logFC=0.8, p-value=0.3), or the severe and critical group (logFC=0.3, p-value=0.61). This suggests MANF is down-regulated in COVID-19 and is not associated with disease severity.

## Discussion

This study explored the changes in protein expression between four disease groups where patients were either controls (tested negative for COVID-19 using PCR) or had mild, severe or critical symptoms of COVID-19. In addition, longitudinal data were available where patients had blood drawn at two different time points after the onset of disease symptoms. In total nine DE analysis were performed (“control vs mild”, “control vs severe”, “control vs critical”, “mild vs severe”, “mild vs critical”, “severe vs critical”, “control vs case”, “mild group longitudinal” and “critical group longitudinal”) to identify proteins significantly perturbed between various disease groups and within groups across time. Furthermore, the results have been made readily available in an interactive web-based R Shiny application (https://phidatalab-shiny.rosalind.kcl.ac.uk/COVID19/), allowing researchers to swiftly visualize and further investigate the expression changes of specific proteins in COVID-19 patients.

### Biomarkers for COVID-19 infection

The three different symptom severity groups (mild, severe and critical) were merged to create a “case” group and were compared to the control group to identify common differentially expressed proteins in COVID-19. A total of 269 proteins were identified to be significantly differentially expressed, of which 120 are up-regulated, and 149 are down-regulated in COVID-19 cases. Notably, over 75% of proteins measured in this study are significantly perturbed in COVID-19 cases as compared to COVID-19 negative controls of similar age groups. Neurofibromin 2 (NF2) was identified as the most perturbed protein in this study and regardless of disease severity, was significantly down-regulated in all COVID-19 patients (Figure 1). This protein was not perturbed according to the longitudinal analysis, and therefore, is not regarded to be associated with the duration of disease. The NF2 protein, or better known as the Merlin protein (moesin-ezrin-radixin-like protein), functions as a tumour suppressor through impacting mechanisms related to proliferation, apoptosis, survival, motility, adhesion, and invasion (9,10). The Merlin protein also activates anti-mitogenic signalling at tight-junctions; hence, an inactivation of Merlin causes uncontrolled mitogenic signalling and tumorigenesis (11).

### Biomarkers for infection severity

Six proteins (IL6, CKAP4, Gal-9, IL-1ra, LILRB4 and PD-L1) were consistently differentially expressed between the control, mild, severe and critical symptom groups. The expression of all six proteins increased as the severity of the symptoms increased, suggesting these proteins may be a useful tool in monitoring the progression from mild to critical disease. Furthermore, these six proteins are not significantly perturbed within the longitudinal analysis, suggesting these proteins are not associated with the length of the duration of infection.

Interleukin 6 (IL6) is an inflammatory cytokine that is an endogenous pyrogen of inducing fever in patients with autoimmune diseases or infections. IL6 is an acute phase inflammatory cytokine that has been suggested to reflect the inflammatory state of the lungs. Elevated IL6 levels have been discovered in ARDS and lung transplantation complications (4,12), and has already been shown to be elevated in COVID-19 patients (12–14). Furthermore, it has also been suggested to be associated with increased COVID-19 mortality (15). Our results add strong support that IL-6 expression increases with disease severity.

Furthermore, while not assessed in this study, IL6 does show some evidence of decreasing in disease remission in COVID-19 patients (12,16) and is, therefore, a viable target for treating the cytokine storm during disease progression. Tocilizumab is a monoclonal antibody targeted against IL6 and its receptor (IL6R) inhibitor that is commonly used to treat inflammatory and autoimmune conditions (17). It is currently being investigated in its effectiveness to treat COVID-19 patients (18,19).

In addition to IL6, we have highlighted novel protein markers associated with disease severity in COVID-19 patients and are all involved in the immune system, with many mediating cytokine productions, including IL6. Cytoskeleton Associated Protein 4 (CKAP4) is involved in the innate immune system and mediates the anchoring of endoplasmic reticulum to microtubules (20). A recent study identified that serum CKAP4 levels of lung cancer patients were significantly higher than those of healthy controls, suggesting CKAP4 as a potential early biomarker for lung cancer (21). Galectin 9 (Gal-9) belongs to a family of beta-galactoside-binding proteins that are implicated in modulating cell-cell and cell-matrix interactions and has a diverse role in the innate and adaptive immune system [provided by RefSeq, Jul 2008]. Gal-9 has been demonstrated to activate ERL/2 phosphorylation, inducing chemokine and cytokine production, including IL-6 (22,23). Serum Gal-9 concentrations have been observed to be significantly increased in patients with infections such as HIV (24), hepatitis C virus (HCV) (25) and malaria (26), suggesting increased gal-9 production is not specific to COVID-19 infection. Interleukin-1 receptor antagonist protein (IL-1ra) inhibits the activities of interleukin 1 alpha (IL1A) and interleukin 1 beta (IL1B) and modulates a variety of interleukin 1 related immune and inflammatory responses [provided by RefSeq, Jan 2016]. Essentially, IL-1ra is an inflammation-inhibitor protein that has also been identified to be significantly higher in COVID-19 patients with a severe symptom (27). Anakinra is a recombinant IL-1ra that has already been administered to COVID-19 patients with suggested improved clinical outcome in two small uncontrolled studies (28,29). Randomized controlled studies are still ongoing.

Leukocyte Immunoglobulin Like Receptor B4 (LILRB4) protein belongs to a family of cell surface receptors that have been suggested to down-regulate the immune response by Inhibiting monocyte activation and inhibiting the production of a critical pro-inflammatory cytokine (TNFα) (30). Increased expression of LILRB4 was associated with increased disease severity in this study, suggesting a possible decrease in monocyte activation leading to an immune-suppressive microenvironment. LILRB4 represents a compelling target to investigate COVID-19 treatment. Programmed cell death 1 ligand 1 (PD-L1) is a type 1 transmembrane protein that has immunoglobulin domains which bind to receptors commonly found on T-cells to inhibit T-cell activation and cytokine production. During infection or inflammation of healthy tissue, this interaction is essential for preventing autoimmunity by maintaining homeostasis of the immune response [provided by RefSeq, Sep 2015]. PD-L1 is found in higher concentrations on some types of cancer cells than healthy cells, which, when bound to PD-1 on T-cells, prevents the T cell from killing the PD-L1 containing cancer cell. To address this, immune checkpoint inhibitors (ICIs) are commonly used in various cancers to block PD-L1 on the cancer cell binding to the T cell, reinvigorating antitumor immune responses (31). However, this immunodeficiency in cancer patients may be the primary cause of why they represent a vulnerable population in the COVID-19 pandemic (32). As these novel proteins are not associated with the duration of infection in neither the mild nor the severe group, they may hold potential as biomarkers for disease severity. However, as members of the immune system, it is unknown if these proteins are markers of general infection rather than COVID-19, thus; they require further investigation for disease specificity, but together, may still be valuable as an additional biomarker for COVID-19 severity after disease confirmation.

### Biomarkers for infection duration

In this study, six patients from the mild group and five patients from the critical group were sampled at two different time-points to identify biomarkers for disease duration. This identified thirteen (BOC, KYNU, SPRY2, KIM1, SCF, MANF, SLAMF1, CD84, SCF, PADI2, PAPPA, CLEC4A, TANK) and six (DECR1, TPSAB1, TF, GDF-8, GZMA, BCAN) proteins in the mild and the critical group respectively, where proteins expression significantly changed from the baseline sample to the first repeat. No protein was discovered to be significantly perturbed in both groups over time; therefore, these biomarkers may be specific within their respective symptom severity groups. However, it is essential to note that sampling time between the baseline and repeat sample differed between the two severity groups, with the mild group averaging 16.2 days and the critical group patients averaging only 2.8 days. The smaller number of days between repeat sampling in the critical symptom group may have been inadequate to measure significant changes in protein expression that reflect infection duration.

### CNS injury biomarkers

We have previously shown neurochemical evidence of neuronal injury and glial response in patients with severe and critical COVID-19. The results of this study indicate that astrocytic activation and/or injury (GFAP) may be a common feature in mild and severe stages of COVID-19, while neuronal injury (NfL) occurs later in the disease process and mainly in patients with critical symptoms (8). In this current study, we expanded our proteomic profiling of CNS proteins, on the same patients, by using the OLINK neurology panel. Correlation analysis identified SCARB2 is most correlated with both tau and GFAP, and EDA2R is most correlated with Nfl, suggesting that these novel proteins are associated with COVID-19-related early or later CNS injury, respectively. Furthermore, MANF and LAT were significantly down-regulated in COVID-19 cases as compared to controls, with expression patterns suggesting it is not associated with disease severity. Although MANF and LAT were present on the OLINK neuronal panel, both proteins are not specific to the brain. Mesencephalic Astrocyte Derived Neurotrophic Factor (MANF) is an endoplasmic reticulum (ER) stress protein and has been suggested to have neuroprotective effects against cerebral ischemia (33). Linker For Activation Of T Cells (LAT) protein is primarily expressed in T-cells and is required for T-cell antigen receptor (TCR) and pre-TCR-mediated signalling. It is important to note that our putative biomarkers of disease progression in COVID-19 (IL6, CKAP4, Gal-9, IL-1ra, LILRB4 and PD-L1) were associated to NfL but to a much lesser extent to GFAP and tau.

The CNS involvement in COVID-19 is not known, and direct invasion of the virus may be unlikely. Support for a hypothesis of CNS infection through a nasopharyngeal route is provided by clinical observations of frequent and persistent anosmia dysgeusia (16). Neurological symptoms are reported in severe cases, supporting the concept that CNS symptoms might be secondary to severe respiratory failure (34). CNS hypoxia from respiratory failure caused by COVID-19, thrombotic microangiopathy, or an indirect effect of extensive cytokine activation that is commonly found in severe COVID-19 is more probable explanations of these increases (Kanberg et al., 2020). Thus, this explorative study of the neurology-associated proteins in the blood may add further insight into hypoxia biomarkers in a broader sense, e.g., to monitor cardiac arrest. Furthermore, the evidence that specific CNS proteins are detectable in blood, as shown by this study, may open up avenues of investigation in CNS injury for neurodegenerative disorders, multiple sclerosis or HIV, where GFAP and NfL are regularly investigated.

### Limitations

Due to blood samples being taken as part of a routine hospital procedure during an unprecedented time, this study was restricted to a small cohort, certain aspects of this study design were uncontrollable and valuable information was unobtainable. The number of “days since symptom onset”, was identified to be significantly higher in the mild symptom group when compared to the severe and the critical symptom group. Essentially, following the onset of symptoms, samples were drawn from the mild symptom group at a much later date when compared to the severe and the critical symptom group. As a result, expression changes involving the mild group (“mild vs severe” and “mild vs critical”) may reflect the duration a patient has been infected with COVID-19 rather than being a reflection of symptom severity. However, as the longitudinal analysis in this study measures protein expression changes during infection, these results were used to differentiate between expression changes likely due to disease severity and duration of infection.

Furthermore, information on comorbidities, medical history and medications are unknown. Hospitalized patients with COVID-19 are known to be more likely to have an underlying health disorder such as hypertension, obesity and diabetes (35), and it is unknown if this cohort has the same characteristics. Moreover, it is unknown if any medication, in addition to oxygen supplementation, was administered to COVID-19 patients, therefore; protein expression changes in this study may be a reflection of a combination of COVID-19, comorbidity and medication.

## Conclusion

This extensive proteomic analysis unbiasedly identified IL6 and five novel proteins (CKAP4, Gal-9, IL-1ra, LILRB4 and PD-L1) to be associated with disease severity in COVID-19 cases. Elevated levels of proteins significantly increased as the disease symptom severity deteriorated and highlight a shared mechanism of cytokine-mediated lung injury cause by viral injection. These proteins warrant further investigation but could provide potential as early biomarkers for disease severity and may serve as potential therapeutic targets, or as biomarkers to monitor the effect of treatments to modulate the immune system and/or suppress the infection. Overall, the results of this study further increase the understanding of COVID-19, which includes CNS involvement, and have been made widely available to researchers as an interactive web-based tool accessible at https://phidatalab-shiny.rosalind.kcl.ac.uk/COVID19/.

## Materials and Methods

### Cohort

Fifty-nine patients with confirmed COVID-19 and 28 healthy, age-matched controls were included. Samples were collected at diagnosis and repeated when possible. Patients were divided into three groups related to systemic disease severity: 26 patients with mild (i.e., not requiring hospitalization) 9 with severe (hospitalized and requiring oxygen supplementation), and 24 with critical disease (admitted to intensive care unit [ICU] and placed on mechanical ventilation [n=23] or not considered a candidate for ICU treatment and with fatal outcome [n=1]). The controls were initially recruited as cognitively unimpaired controls for an observational dementia study, and they were all neurologically and psychiatrically normal with any magnetic resonance imaging (MRI) abnormalities set as an exclusion criteria. Follow-up samples on patients with critical COVID-19 were collected when they were still in ICU.

### COVID-19 confirmation

The diagnosis was confirmed using real-time polymerase chain reaction (rtPCR) analysis of nasal and throat swab specimens. Nucleic acid was extracted from clinical samples in a MagNA Pure 96 instrument using the Total Nucleic Acid isolation kit (Roche). rtPCR targeting the RdRP region was performed in a QuantStudio 6 instrument (Applied Biosystems, Foster City, CA) using the probe described by Corman et al. and the primers RdRP_Fi, GTCATGTGTGGCGGTTCACT and RdRP_Ri, CAACACTATTAGCATAAGCAGTTGT (36).

### Plasma proteomics

Plasma glial fibrillary acidic protein (GFAP), neurofilament light (NfL), and tau were measured and reported in our recent COVID-19 related study (8), which discovered neurochemical evidence to support the possible impact of COVID-19 on the CNS. The Simoa protein measurements are detailed in (8). Plasma from the same participants was also used for the OLINK protein profiling for this study. Protein concentrations were measured on the Olink Multiplex platform (Olink Proteomics AB, Uppsala, Sweden) using the cardiovascular II (v.5006), immune response (v.3203), inflammation (v.3022) and neurology (v.8012) 96-plex panels. The OLINK immunoassays are based on the Proximity Extension Assay (PEA) technology (37), which uses a pair of oligonucleotide-labelled antibodies to bind to their respective target protein. When the two antibodies are in close proximity, a new polymerase chain reaction target sequence is formed, which is then detected and quantified by quantitative real-time PCR.

### Statistical analysis

The Olink-generated data was preprocessed and quality controlled using the platform-specific “Olink NPX manager” software, which background corrects, log2 transforms and normalizes all samples to an arbitrary NPX scale. The NPX is a relative quantification unit where a difference of 1 NPX equates to a doubling of protein concentration.

Additional data processing was performed in RStudio (version 1.2.1335) using R (version 3.6.0). First, samples with a failure rate of more than 50% across all proteins were removed. Next, proteins were removed if the protein failed to quantify in more than 50% of samples in each disease group, or if the protein NPX value fell below the protein-specific LOD value in more than 50% of samples in each disease group. The remaining NPX values below the LOD were substituted by the proteins LOD/√2.

The demographic variables available were ethnicity, age, gender and “days since symptom onset”. The “days since symptom onset” variable represents the number of days that a blood sample was taken after the first self-reported symptom date. The Welch Two Sample t-test and the Fisher’s exact test was performed where appropriate to identify any significant differences in age, gender and “days since symptom onset” between groups.

DE analysis was performed using the R package “limma” (version 3.42.2) using gender, age and “days since symptom onset” as covariates where possible. A protein was determined to be significantly differentially expressed if the false discovery rate (FDR) adjusted p-value was ≤ 0.05. The following self-explanatory comparisons were made: 1) “control vs mild”, 2) “control vs severe”, 3) “control vs critical”, 4) “mild vs severe”, 5) “mild vs critical”, 6) “severe vs critical”, 7) “control vs case” (where patients with mild, severe and critical symptoms are merged and treated as the cases).

Some patients within the mild and the severe symptom group had protein concentrations measured at two different time points; therefore, two additional DE analysis was performed independently in the mild and the severe symptom groups and are referred to as 1) “mild group longitudinal” and 2) “critical group longitudinal” analysis, respectively. The two longitudinal analyses were performed in their respective disease groups using a paired t-test approach in limma.

Pathway enrichment analysis was performed using an Over-Representation Analysis (ORA) implemented through the ConsensusPathDB (http://cpdb.molgen.mpg.de) web-based platform (version 34) (38). Significant results were then explored using the “KEGG mapper – Search&Color Pathway” to map and visualize proteins in a specific biological pathway (39).

This study further explores CNS injury-related biomarkers in COVID-19 by correlating the Simoa measured proteins with the OLINK measured proteins using Pearson’s correlation.

## Data availability

The proteomic data is available in the BioStudies database (http://www.ebi.ac.uk/biostudies) under accession number S-BSST416. Additionally, an R shiny application was written in R using the “shiny” framework (version 1.4.0.2) to allow the quick and efficient visualization of the expression of specific proteins across the control, mild, severe and severe symptom groups. The application is hosted on the research computing facility at King’s College London (Rosalind), and allows researchers to quickly visualize and investigate the results across all nine DE analyses performed in this study. The application can be accessed at https://phidatalab-shiny.rosalind.kcl.ac.uk/COVID19/. All data analysis scripts used in this study have been deposited in zenodo under the DOI: 10.5281/zenodo.3895886

## Ethics Statement

This study has been approved by the Swedish Ethical Review Authority (2020-01771). All participants provided written informed consent, in those with severe COVID-19, this was obtained before they were placed on mechanical ventilation and were deemed fully capable of understanding the nature of the study and their part in.

## Acknowledgements

This study presents independent research supported by the NIHR BioResource Centre Maudsley at South London and Maudsley NHS Foundation Trust (SLaM) & Institute of Psychiatry, Psychology and Neuroscience (IoPPN), King’s College London. The views expressed are those of the author(s) and not necessarily those of the NHS, NIHR, Department of Health or King’s College London. The authors acknowledge use of the research computing facility at King’s College London, Rosalind (https://rosalind.kcl.ac.uk), which is delivered in partnership with the National Institute for Health Research (NIHR) Biomedical Research Centres at South London & Maudsley and Guy’s & St. Thomas’ NHS Foundation Trusts, and part-funded by capital equipment grants from the Maudsley Charity (award 980) and Guy’s & St. Thomas’ Charity (TR130505). The views expressed are those of the author(s) and not necessarily those of the NHS, the NIHR, King’s College London, or the Department of Health and Social Care.

NJA is supported by the Wallenberg Centre for Molecular and Translational Medicine, the Swedish Alzheimer Foundation (Alzheimerfonden), the Swedish Dementia Foundation (Demensförbundet), Hjärnfonden, Sweden and the Anna Lisa and Brother Björnsson’s Foundation. RJBD is supported by 1. Health Data Research UK, which is funded by the UK Medical Research Council, Engineering and Physical Sciences Research Council, Economic and Social Research Council, Department of Health and Social Care (England), Chief Scientist Office of the Scottish Government Health and Social Care Directorates, Health and Social Care Research and Development Division (Welsh Government), Public Health Agency (Northern Ireland), British Heart Foundation and Wellcome Trust. 2. The National Institute for Health Research University College London Hospitals Biomedical Research Centre. KB is supported by the Swedish Research Council (#2017-00915), the Alzheimer Drug Discovery Foundation (ADDF), USA (#RDAPB-201809-2016615), the Swedish Alzheimer Foundation (#AF-742881), Hjärnfonden, Sweden (#FO2017-0243), the Swedish state under the agreement between the Swedish government and the County Councils, the ALF-agreement (#ALFGBG-715986), and European Union Joint Program for Neurodegenerative Disorders (JPND2019-466-236). MG is supported by The Swedish State Support for Clinical Research (ALFGBG-717531) and by SciLifeLab/KAW national COVID-19 research program project grant (V-2020-0250). HZ is a Wallenberg Scholar supported by grants from the Swedish Research Council (#2018-02532), the European Research Council (#681712), Swedish State Support for Clinical Research (#ALFGBG-720931), the Alzheimer Drug Discovery Foundation (ADDF), USA (#201809-2016862), and the UK Dementia Research Institute at UCL.

## Conflicts of interest

KB has served as a consultant, at advisory boards, or at data monitoring committees for Abcam, Axon, Biogen, Julius Clinical, Lilly, MagQu, Novartis, Roche Diagnostics, and Siemens Healthineers, and is a co-founder of Brain Biomarker Solutions in Gothenburg AB (BBS), which is a part of the GU Ventures Incubator Program (outside submitted work). HZ has served at scientific advisory boards for Denali, Roche Diagnostics, Wave, Samumed, Siemens Healthineers, Pinteon Therapeutics and CogRx, has given lectures in symposia sponsored by Fujirebio, Alzecure and Biogen, and is a co-founder of Brain Biomarker Solutions in Gothenburg AB (BBS), which is a part of the GU Ventures Incubator Program (outside submitted work).

## Supporting information captions

Supplementary Table 1: Statisitcal testing of patient demographic variables

Supplementary Table 2: Complete differential expression analysis results

Supplementary Table 3: Pathway enrichment analysis results

Supplementary Table 4: OLINK neuronal proteins correlated with markers of neural injury and astrogliosis

